# Buprenorphine treatment receipt characteristics and retention among people who inject drugs at Integrated Care Centers in India

**DOI:** 10.1101/2022.08.29.22279348

**Authors:** Lakshmi Ganapathi, Allison M. McFall, Kimberly F. Greco, Aylur Srikrishnan, Kenneth H. Mayer, Conall O’Cleirigh, Shruti H. Mehta, Gregory M. Lucas, Sunil S. Solomon

**Affiliations:** Division of Infectious Diseases, Boston Children’s Hospital, Boston, Massachusetts; Department of Pediatrics, Boston Children’s Hospital, Boston, Massachusetts; Harvard Medical School, Boston, Massachusetts; Department of Epidemiology, The Johns Hopkins Bloomberg School of Public Health, Baltimore, Maryland; Institutional Centers for Clinical and Translational Research, Boston Children’s Hospital, Boston, Massachusetts; YR Gaitonde Centre for AIDS Research and Education, Chennai, India; Division of Infectious Diseases, Beth Israel Deaconess Medical Center, Boston, Massachusetts; The Fenway Institute, Boston, Massachusetts; Department of Psychiatry, Massachusetts General Hospital, Boston, Massachusetts; Division of Infectious Diseases, The Johns Hopkins University School of Medicine, Baltimore, Maryland

**Keywords:** people who inject drugs, opioid use, buprenorphine, treatment, retention, India

## Abstract

**Background:** India is facing an alarming rise in the injection of opioids leading to burgeoning HIV epidemics among people who inject drugs (PWID) in several cities. Integrated Care Centers (ICCs) provide free single-venue HIV services and substance use treatment to PWID and have been established across 8 Indian cities. We evaluated engagement of PWID in buprenorphine treatment at ICCs to inform interventions.

**Methods:** We retrospectively analyzed 1-year follow-up data for PWID initiating buprenorphine between 1 January – 31 December 2018 across 7 ICCs. We used descriptive statistics to evaluate buprenorphine uptake, receipt frequency, treatment interruptions (no buprenorphine receipt for 60 consecutive days but with subsequent re-engagement in treatment), and treatment drop-out (no buprenorphine receipt for 60 consecutive days without subsequent re-engagement), and explore differences between historical opioid epidemic regions (i.e., Northeast cities (NEC)) and emerging opioid epidemic regions (i.e., North/Central/Northwest cities (NCC)). We used a multivariable logistic regression model to determine predictors of treatment drop-out by 6 months.

**Results:** 1312 PWID initiated buprenorphine (76% NCC vs. 24% NEC). 31% of PWID in NCC, and 25% in NEC experienced ≥ 1 treatment interruption. About a third (34%) of PWID in NCC vs. half (50%) in NEC dropped-out by 6 months (p<0.0001). Over 6 months, 48% of PWID in NCC vs. 60% in NEC received buprenorphine ≤2 times/week on average (p<0.0001). In multivariable models, living in NEC was associated with increased odds of treatment drop-out while receipt of counseling was associated with decreased odds of treatment drop-out.

**Conclusions:** PWID at ICCs, particularly those in NEC have low buprenorphine receipt and retention. Patient-centered interventions adapted to regional contexts are urgently needed to ameliorate these gaps.

## 1.1 Introduction

India has among the largest number of opioid users in the world (∼23 million)^1,2^. Opioid use (primarily heroin, pharmaceutical opioids and opium and opium variants) is shaped by several factors. Opium cultivation is legal in selected states^3^. Given India’s location between regions of heroin production in Southeast Asia and Central Asia, its Northeast and Northwest states are both destinations and intermediary routes of heroin trafficking. Additionally, large-scale diversion of pharmaceutical opioids within and outside the country occurs in the context of a thriving pharmaceutical industry and poor regulation of pharmacies and legal opioid production^4^.

National estimates suggest that at least 10 million Indians have opioid dependence and/or engage in harmful use^1^. Scale up of treatment for opioid dependence in the public sector (known as “Opioid Substitution Therapy” or OST in local nomenclature which is retained in this manuscript) initially via provision of buprenorphine, and buprenorphine-naloxone and subsequently with the addition of methadone in some states has occurred over three decades in India^5,6^. However, OST availability commensurate with the magnitude of need has been inadequate and treatment has largely been limited to people who inject drugs (PWID)—the vast majority of whom inject opioids – to address the HIV and viral hepatitis epidemics in this population^5,7^.

Public sector services to address opioid use disorders and substance use more broadly have primarily relied on provision of inpatient treatment at government hospitals^8^. Systematic introduction of substance use treatment to PWID at community-based venues began between 2007 – 2012, when as part of the National AIDS Control Program, The National AIDS Control Organization (NACO) established the “OST Program”^5,6^. The OST program initially comprised 52 OST centers run by non-governmental organizations (NGOs) at community locations. These OST centers provided buprenorphine via daily observed therapy to PWID (a policy for medication dispensing that is still in place). To expand the OST program’s reach, NACO subsequently established a collaborative model between government hospitals and more than 150 NGOs providing “targeted interventions” (i.e., other harm reduction services such as needle and syringe exchange, condoms etc.) to PWID. The hospitals housed OST centers, with a medical team (a doctor, nurse, and counselor) responsible for assessments, daily dispensing of buprenorphine/ methadone, and clinical follow-up while partner NGOs continued to provide other harm reduction services, conducted outreach, provided initial referrals to the OST centers, and undertook subsequent community-based follow-up of PWID. Limitations of this model included fragmentation of services between multiple physical locations that increased barriers for receipt of OST and other HIV services among PWID.

Over the last decade, there has been rapidly expanding injection drug use epidemics (mainly with pharmaceutical opioids) fueling HIV and hepatitis C epidemics in multiple states in the North/Central/Northwest regions of India which have a paucity of services for PWID^9–16^. To address this dearth and circumvent fragmentation in prior service delivery models, since 2013, “Integrated Care Centers” (ICCs) – a public-private service delivery model between NACO, State AIDS Control Societies and NGOs – have been established across 8 cities in India spanning the Northeast and North/Central/Northwest regions^17^. ICCs are situated in community locations and offer single-venue co-located outpatient services to PWID that includes provision of OST, HIV and hepatitis C testing, anti-retroviral therapy (ART) and hepatitis C treatment on-site or via linkage to government centers, needle and syringe exchange, condoms, tuberculosis screening, and counseling services. Physicians, nurses, and counselors are on-site at ICCs to provide the clinical services. Additionally, peer educators and other program staff are involved in outreach and community follow up. The vast majority of PWID (>95%) at ICCs receive buprenorphine, although select ICCs also provide methadone.

We undertook this retrospective study to obtain a global view of buprenorphine receipt among PWID who visit ICCs to receive one or more services, and to specifically examine treatment receipt characteristics and predictors of treatment drop-out among PWID newly initiating buprenorphine treatment. This study additionally sought to characterize regional differences in treatment receipt and delineate patient-level and program-level gaps.

## 1.2 Materials and Methods

### 1.2.1 Setting

There are currently 7 ICCs in operation: three in the Northeast cities (NEC) of Aizawl (Mizoram State), Churchandpur (Manipur State), and Dimapur (Nagaland State) and five in the North/Central/Northwest cities (NCC) of Ludhiana and Amritsar (Punjab State), Bilaspur (Chhattisgarh State), Kanpur (Uttar Pradesh State), and New Delhi. PWID receiving services at ICCs receive a client identification number at registration which is linked to unique biometric fingerprinting data. Visits to receive any services including OST are captured via this biometric verified linked ID. At registration, demographic data are collected, and all PWID receive HIV and hepatitis C testing with reminders every 6 months to repeat testing. Counseling services provided at ICCs include pre- and post-test HIV counseling as well as substance use-specific counseling comprising brief interventions such as motivational interviewing. Sociodemographic data, and visits to receive services, including referrals out of ICCs for additional services (e.g., for tuberculosis treatment) are recorded and stored in a centralized database, linked to client identification numbers.

### 1.2.2 Data extraction and definitions

Given the COVID-19 pandemic’s impact on how OST was delivered in the ICCs between 2020-2021 and the limited ability to accurately track OST visits in the database, for this study, we used pre-pandemic data. To characterize receipt of buprenorphine among *all* PWID receiving services at ICCs, we included those who had registered at an ICC and had at least one visit for any service between January 1, 2018, and December 31, 2018. To specifically examine treatment receipt characteristics and treatment drop-out among PWID who newly initiate buprenorphine, we assembled a cohort of PWID who initiated buprenorphine between January 1, 2018, and December 31, 2018. Entry into the cohort occurred when a PWID first registered to receive OST. We examined 1 year follow-up data following initiation for all PWID in the cohort (i.e., last possible follow-up through December 31, 2019) to determine treatment interruptions and treatment drop-out. A treatment interruption was defined as not presenting to receive buprenorphine for 60 consecutive days but subsequently re-engaging in treatment within the study period. Treatment drop-out was defined as not presenting to receive buprenorphine for 60 consecutive days without subsequent re-engagement in treatment for the duration of the study period. While there are no universally accepted definitions for treatment interruptions and treatment drop-out, we chose 60 days in alignment with previous studies among people receiving medications for opioid use disorders that have defined treatment interruptions and/ or treatment drop-out based on consecutive non-attendance for time periods anywhere between 30 and 90 days^18–21^. We additionally examined buprenorphine receipt frequency on average over the course of the first 6 months. We chose treatment drop-out by 6 months as an outcome for additional analysis for the following reasons: NACO’s a priori programmatic benchmarks for OST receipt at ICCs include at least 50% retention in OST at 6 months; Treatment drop-out by 6 months was also chosen to compare these outcome metrics at ICCs with other similar studies in India and globally. As Kanpur’s ICC only started providing OST in 2019, it was excluded from analysis which comprised data from the other 7 ICCs.

### 1.2.3 Statistical analysis

To characterize buprenorphine receipt, we generated overall descriptive data for ICCs, and further stratified these data by region (Northeast ICCs versus North/Central/Northwest ICCs). This regional stratification was chosen for several reasons. The Northeast cities represent historical opioid injection epidemics whereas the North/Central/Northwest cities represent emerging opioid injection epidemics^22–24^. Consequently, there have been longer standing harm reduction initiatives in the Northeast cities including the delivery of OST via NGOs as compared to fewer if any pre-existing initiatives prior to the establishment of ICCs in the North/Central/Northwest cities. To determine regional differences in treatment receipt characteristics among PWID newly initiating buprenorphine, we used chi-square tests for categorical variables and 2-sample Wilcoxon tests for continuous variables. We generated a histogram for maintenance dose of buprenorphine received in the first 6 months following initiation for the cohort, and separately classified frequencies for the following dose categories: < 4mg, 4-7 mg, 8-12 mg and >12 mg. These dose categories were based on Indian OST national guidelines that recommend 8-12 mg as generally optimal maintenance dosing^25^. To determine predictors of treatment drop-out by 6 months, we used a multivariable logistic regression model. Predictors included sociodemographic characteristics (age, gender, marital status, employment, income), HIV status, region, receipt of counseling and maintenance buprenorphine dose. All variables assessed in univariable analysis were included in the multivariable model based on prior evidence of likely association with treatment drop-out^18,26– 31^. For significant variables identified in the multivariable models, we additionally constructed Kaplan Meir survival curves evaluating retention over the course of 1 year. Finally, we conducted sensitivity analyses to determine if more proximate definitions of treatment interruption (non-attendance over 30 days vs. 60 days) and drop-out (loss to follow up by 3 months vs. 6 months) materially affected results. All analyses were conducted using SAS 9.4 (SAS Institute, Cary, North Carolina).

### 1.2.3 Ethics statement

This study utilized de-identified and de-linked data. The study received ethics approval from the Institutional Review Boards of The Johns Hopkins University School of Medicine, Boston Children’s Hospital and the Y.R. Gaitonde Centre for AIDS Research and Education.

## 1.3 Results

### 1.3.1 Buprenorphine receipt among PWID at ICCs

In 2018, 5148 unique PWID received at least one service (Table 1) across 7 ICCs (NEC: 1092 PWID, NCC: 4056 PWID). Approximately three quarter (74%) were new registrants to the ICC (NEC: 71%, NCC: 75%). Among all PWID receiving services at ICCs, overall, 60% received buprenorphine at least once (NEC: 56%, NCC: 61%). Among those receiving buprenorphine, 1312 PWID (43%) newly initiated treatment in 2018 (NEC: 52%, NCC: 40%).

**Table 1.**
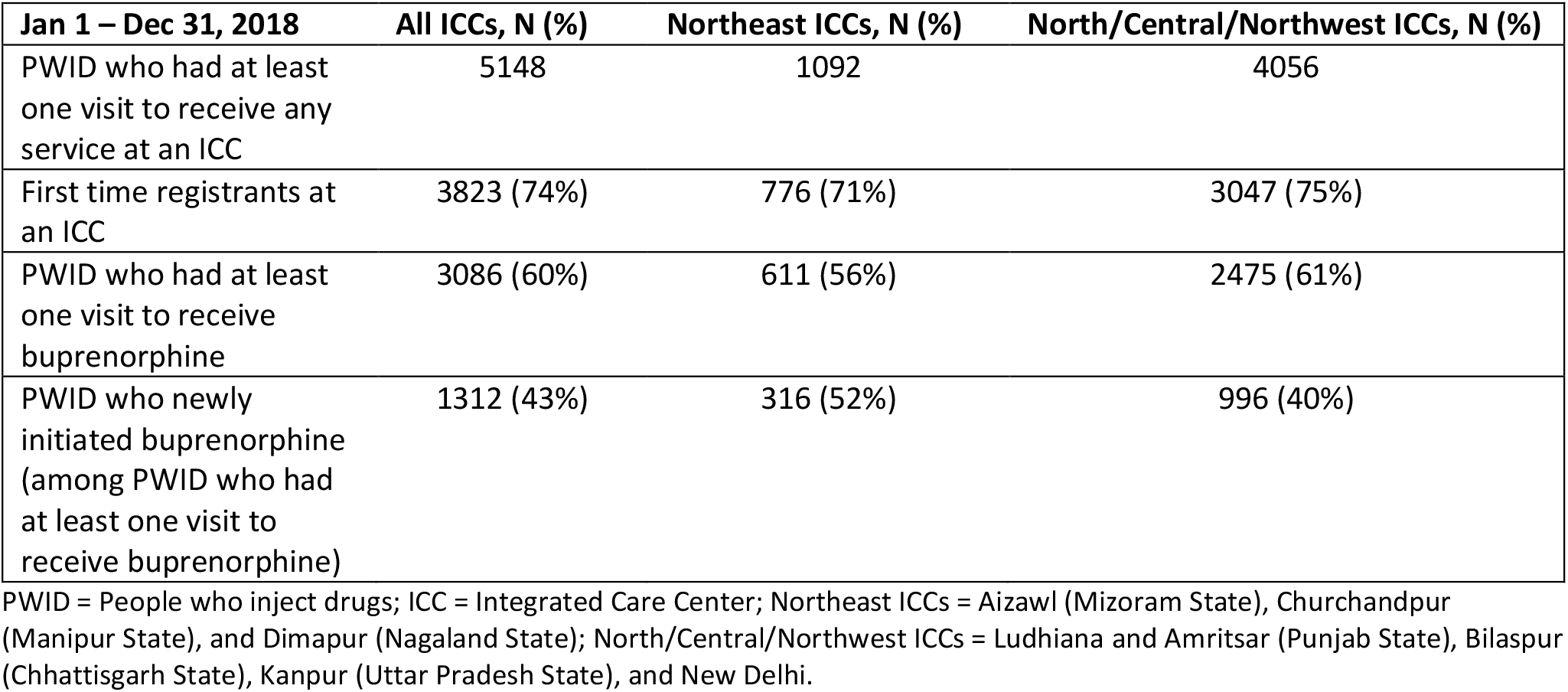
Buprenorphine receipt among people who inject drugs receiving services at Integrated Care Centers in India.

| **Jan 1 – Dec 31, 2018** | **All ICCs, N (%)** | **Northeast ICCs, N (%)** | **North/Central/Northwest ICCs, N (%)** |
| --- | --- | --- | --- |
| PWID who had at least one visit to receive any service at an ICC | 5148 | 1092 | 4056 |
| First time registrants at an ICC | 3823 (74%) | 776 (71%) | 3047 (75%) |
| PWID who had at least one visit to receive buprenorphine | 3086 (60%) | 611 (56%) | 2475 (61%) |
| PWID who newly initiated buprenorphine (among PWID who had at least one visit to receive buprenorphine) | 1312 (43%) | 316 (52%) | 996 (40%) |

PWID = People who inject drugs; ICC = Integrated Care Center; Northeast ICCs = Aizawl (Mizoram State), Churchandpur (Manipur State), and Dimapur (Nagaland State); North/Central/Northwest ICCs = Ludhiana and Amritsar (Punjab State), Bilaspur (Chhattisgarh State), Kanpur (Uttar Pradesh State), and New Delhi.

### 1.3.2 Demographic characteristics of PWID newly initiating buprenorphine

The median age of PWID newly initiating buprenorphine was 27 years (Table 2). In keeping with the gender composition of PWID accessing services at ICCs, more than 98% of PWID who initiated buprenorphine were male. The vast majority of PWID had high school and above education. Approximately 90% of PWID were HIV uninfected at the time of buprenorphine initiation while 8% were HIV infected. Overall, about a third (30%) of PWID were unemployed, although there were observable regional differences. More than 75% of PWID in the Northeast ICCs were unemployed compared to approximately 15% of PWID in the North/Central/Northwest ICCs.

**Table 2.**
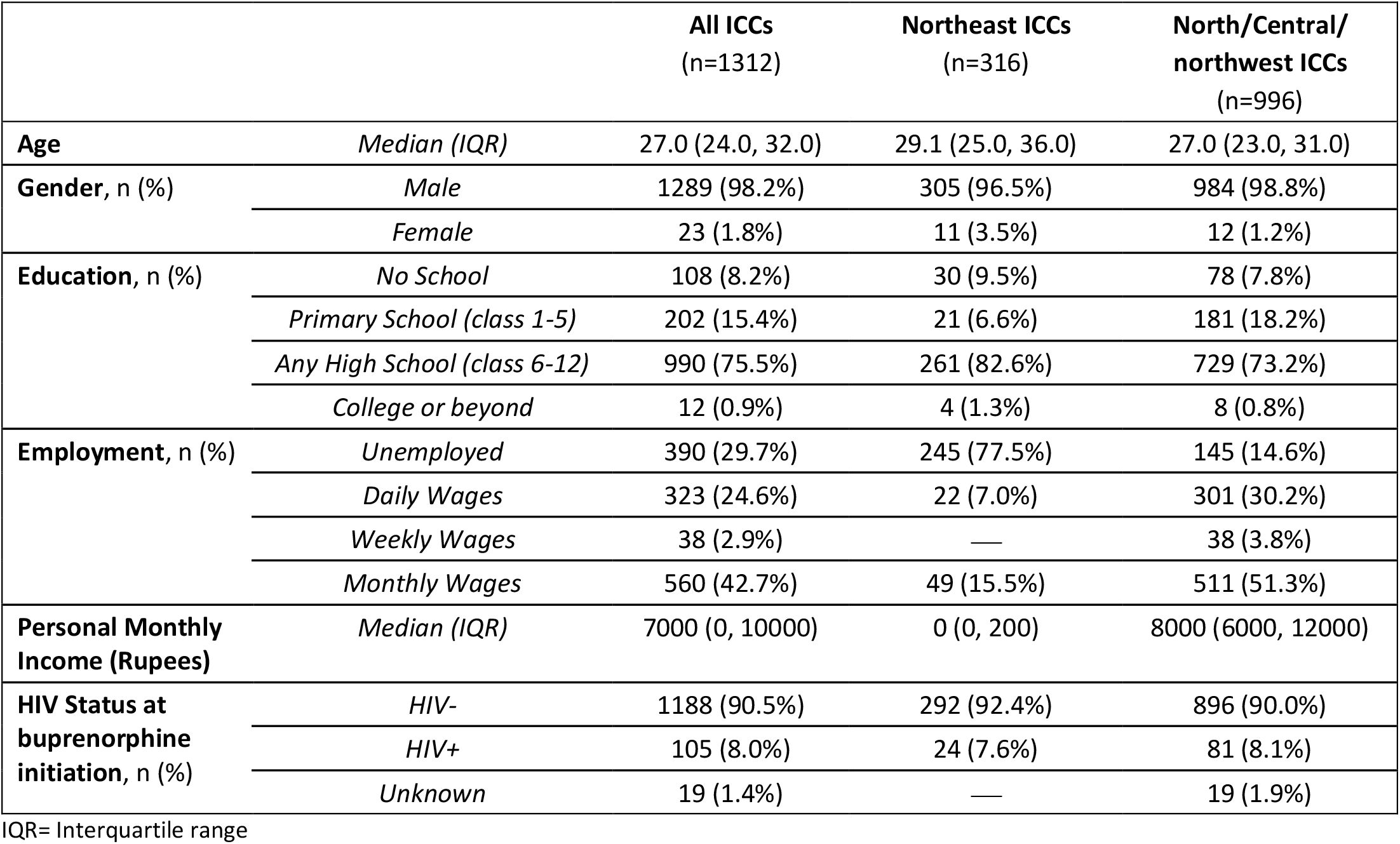
Demographic characteristics of People who Inject Drugs (PWID) who newly initiated buprenorphine at Integrated Care Centers (ICCs) in 2018

### 1.3.3 Treatment receipt characteristics among PWID newly initiating buprenorphine

The median number of days between ICC registration and OST initiation was 1 day (Interquartile [IQR] 0, 3 days). The median number of days buprenorphine was received by PWID in the cohort was 47 days in 6 months, and 62 days in 1 year (Table 3). Approximately 30% experienced at least one treatment interruption. The median number of days to first treatment interruption was 39 days. Approximately 28% of PWID dropped-out of treatment by 3 months, 38% by 6 months, and 65% by one year. Constructing an opioid treatment cascade (Figure 1), 72% of PWID were retained in treatment at 3 months, 62% at 6 months and 35% at 1 year. On average, over the course of 6 months, only about 49% of PWID received buprenorphine at a frequency of more than two times a week. More than 50% did not receive any counseling, or counseling specific to substance use.

**Table 3.** Buprenorphine receipt characteristics among people who inject drugs newly initiating treatment at Integrated Care Centers

|  |  | All ICCs<br>(n=1312) | Northeast ICCs<br>(n=316) | North/Central/<br>northwest ICCs<br>(n=996) | p-value |
| --- | --- | --- | --- | --- | --- |
| **Visits in 6 months (days)** | *Median (IQR)* | 47.0 (6.0, 123.0) | 30.5 (6.0, 84.5) | 57.5 (6.0, 134.0) | <0.0001 |
| **Visits in 1 year (days)** | *Median (IQR)* | 62.0 (8.0, 212.5) | 34.5 (7.0, 119.0) | 78.0 (9.0, 241.5) | <0.0001 |
| **PWID experiencing at least 1 treatment interruption, n (%)** |  | 387 (29.5%) | 80 (25.3%) | 307 (30.8%) | 0.0614 |
| **Days to first treatment interruption\*** | *Median (IQR)* | 39.0 (6.0, 96.0) | 38.0 (9.5, 74.5) | 40.0 (5.0, 101.0) | 0.6630 |
| **Frequency of buprenorphine visits in the first 6 months, n (%)** | *None* | 126 (9.6%) | 33 (10.4%) | 93 (9.3%) | <0.0001 |
|  | *1-3 times/month (≤3/ month)* | 352 (26.8%) | 97 (30.7%) | 255 (25.6%) |  |
|  | *1-2 times/week (4-8/ month)* | 187 (14.3%) | 61 (19.3%) | 126 (12.7%) |  |
|  | *3-5 times/week (9-20/ month)* | 301 (22.9%) | 79 (25.0%) | 222 (22.3%) |  |
|  | *>5 times/week (>20/month)* | 346 (26.4%) | 46 (14.6%) | 300 (30.1%) |  |
| **Frequency of any counseling visits in the first 6 months, n (%)** | *None* | 697 (53.1%) | 126 (39.9%) | 571 (57.3%) | <0.0001 |
|  | *More than once* | 615 (46.9%) | 190 (60.1%) | 425 (42.7%) |  |
| **Frequency of safe injection and substance use counseling visits in the first 6 months, n (%)** | *None* | 766 (58.4%) | 178 (56.3%) | 588 (59.0%) | 0.3950 |
|  | *More than once* | 546 (41.6%) | 138 (43.7%) | 408 (41.0%) |  |
| **Treatment drop-out by 3 months, n (%)** |  | 371 (28.3%) | 119 (37.7%) | 252 (25.3%) | <0.0001 |
| **Treatment drop-out by 6 months, n (%)** |  | 501 (38.2%) | 158 (50.0%) | 343 (34.4%) | <0.0001 |
| **Treatment drop-out by 1 year, n (%)** |  | 857 (65.3%) | 252 (79.7%) | 605 (60.7%) | <0.0001 |
| **Days to treatment drop-out** | *Median (IQR)* | 273.5 (65.5, 365.0) | 179.5 (28.5, 346.5) | 301.5 (87.0, 365.0) | <0.0001 |
| **Treatment drop-out by 6 months among PWID experiencing at least 1 treatment interruption, n (%)\*** |  | 50 (12.9%) | 10 (12.5%) | 40 (13.0%) | 0.9000 |
| **Treatment drop-out by 1 year among PWID experiencing at least 1 treatment interruption, n (%)\*** |  | 282 (72.9%) | 68 (85.0%) | 214 (69.7%) | 0.0061 |

PWID = People who Inject Drugs; IQR = Interquartile range
\* Calculated among n=387 PWID experiencing at least 1 treatment interruption
p-values computed using chi-square analysis for categorical variables and 2-sample Wilcoxon tests for continuous variables

**Figure 1.**
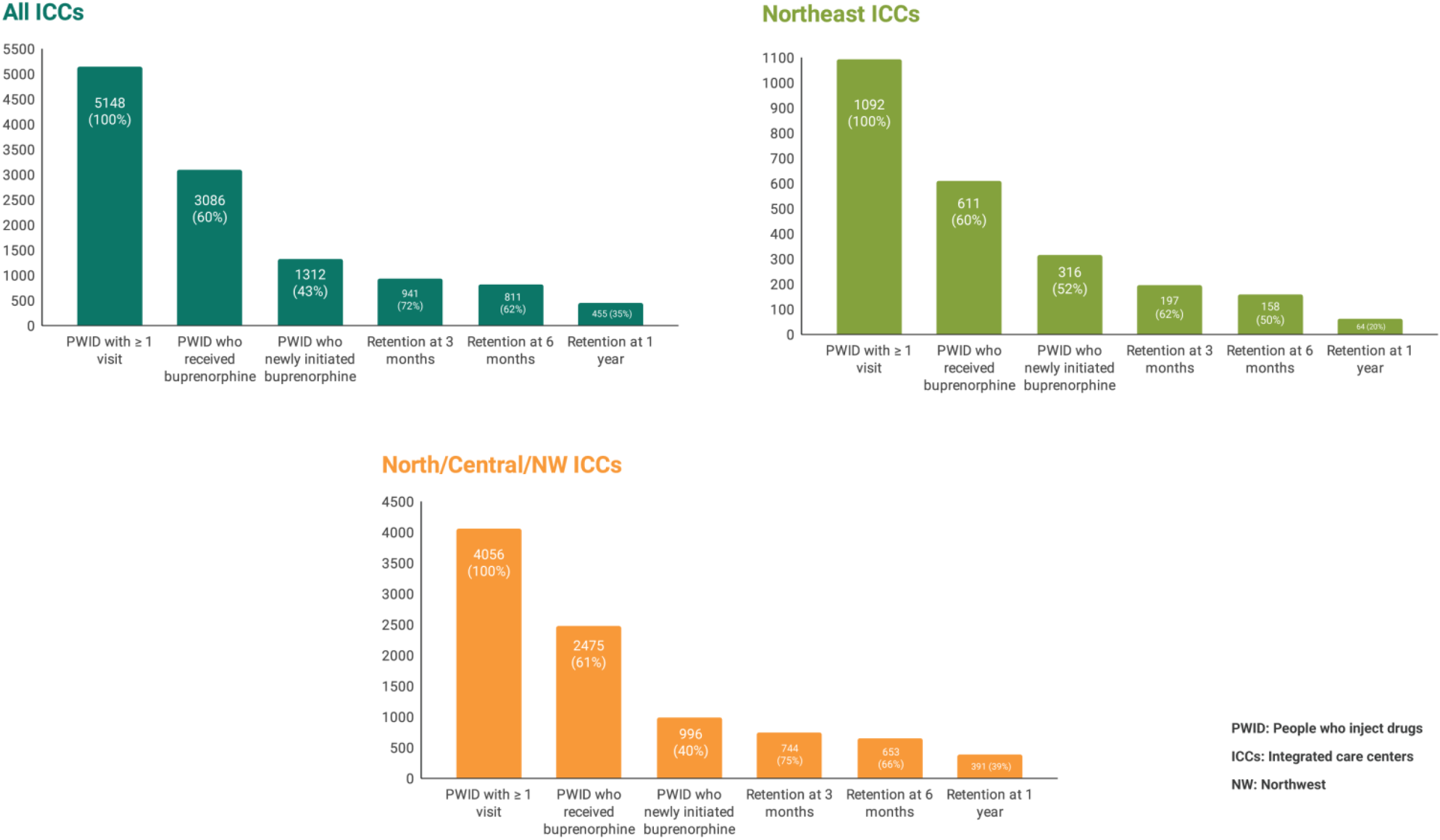
The Opioid Treatment Cascade for People who Inject Drugs receiving Buprenorphine at Integrated Care Centers in India.

There were notable regional differences in treatment receipt characteristics among PWID newly initiating buprenorphine (Table 3). The median number of days of buprenorphine receipt over 6 months and 1 year was significantly lower among PWID in the Northeast ICCs compared to those in the North/Central/Northwest ICCs (NEC: 30 days, NCC: 57 days in 6 months; NEC: 34 days, NCC: 78 days in 1 year, p<0.0001). A significantly greater proportion of PWID at Northeast ICCs dropped-out of treatment by 3 months, 6 months and 1 year respectively. The median time to treatment drop-out was significantly shorter among PWID in the Northeast ICCs compared to those in the North/Central/Northwest ICCs (NEC: 179.5 days, NCC: 301.5 days, p<0.0001). On average, over the course of 6 months, a significantly lower proportion of PWID in the Northeast ICCs received buprenorphine at a frequency of more than two times a week compared to PWID in the North/Central/Northwest ICCs (NEC: 40%, NCC: 52%, p<0.0001).

### 1.3.4 Buprenorphine dose characteristics

Nearly 75% of the cohort did not experience any changes to maintenance buprenorphine dose following initiation. Among the PWID who did have dose changes, 17% had an increase in daily dose while approximately 8% had a decrease in daily dose. We examined the average daily dose prescribed in the 6 months following initiation. The median average daily dose was 6 mg (IQR 4, 6.5 mg). By dose categories (Figure 2), approximately 7% of PWID received buprenorphine dose less than 4 mg, 74% received between 4-7 mg, 14% received between 8-12 mg and 4% received dose greater than 12 mg.

**Figure 2.**
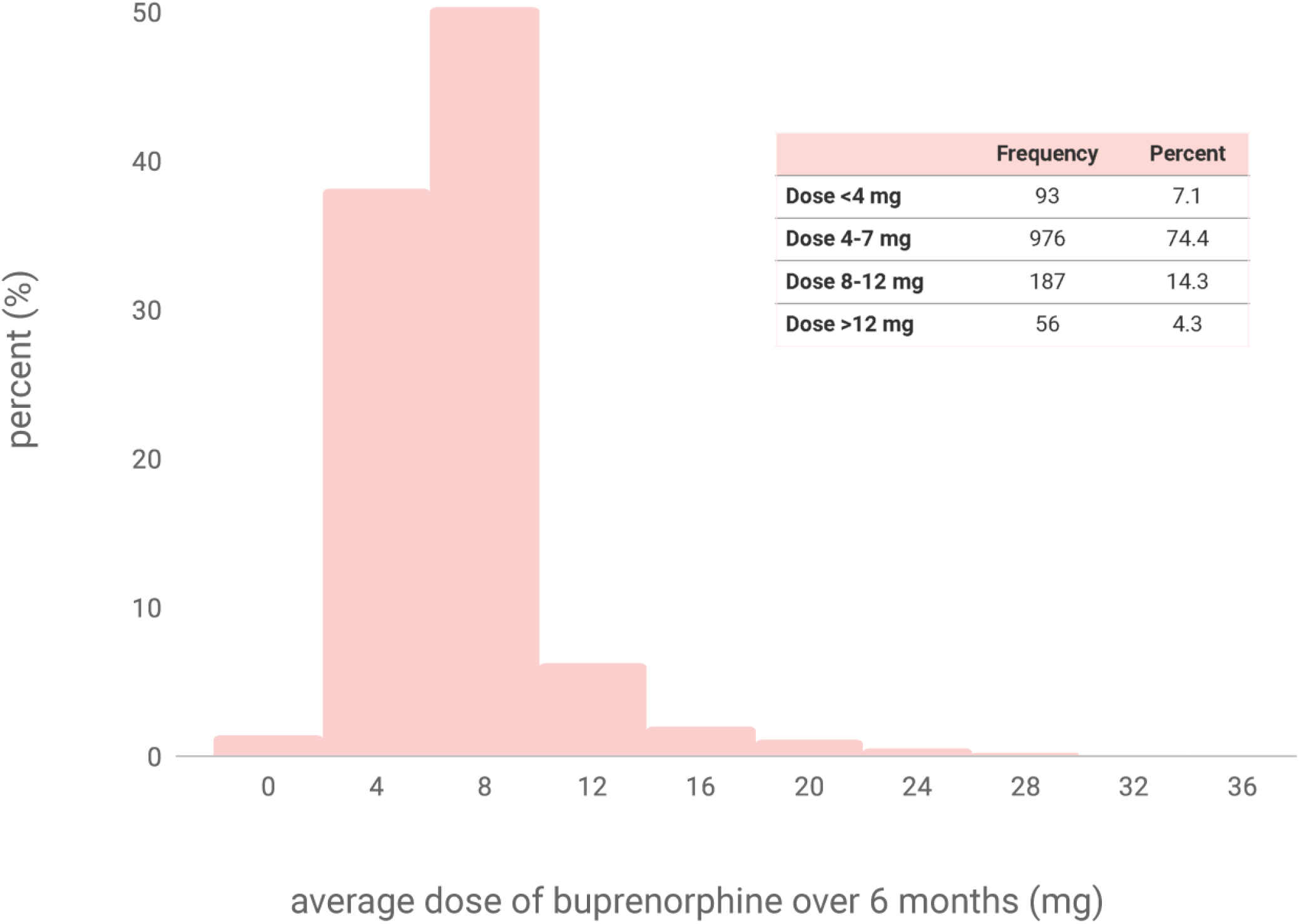
Buprenorphine dose characteristics among people who inject drugs initiating treatment at Integrated Care Centers.

### 1.3.5 Predictors of treatment drop-out by 6 months

In univariable analysis, female PWID, unemployed PWID, and PWID in the Northeast ICCs had higher odds of treatment drop-out by 6 months compared to male PWID, PWID who were employed and those in the North/Central/Northwest ICCs respectively (Table 4). Receipt of any counseling was associated with decreased odds of treatment drop-out by 6 months. In multivariable analysis only region and receipt of counseling were significantly associated with treatment drop-out by 6 months. Notably, PWID in Northeast ICCs had nearly twice the odds of treatment drop-out compared to PWID in the North/Central/Northwest ICCs while PWID who had received any counseling had less than half the odds of treatment drop-out compared to those who had received no counseling. Age, marital status, income, HIV status at OST initiation and maintenance opioid dose were not associated with treatment drop-out by 6 months. Kaplan Meir survival curves for the cohort, and by region and receipt of counseling provides visualization of retention over the 1-year study period and further illuminates differences (Figure 3).

**Table 4.** Predictors of treatment drop-out by 6 months among people who inject drugs initiating buprenorphine at Integrated Care Centers

|  |  | OR (95% CI) | aOR (95% CI)<sup>‡</sup> |
| --- | --- | --- | --- |
| **Age at buprenorphine initiation** | per year | 1.00 (0.98, 1.01) | 1.00 (0.98, 1.02) |
| **Gender** | Female | 2.56 (1.10, 5.97)* | 2.42 (0.94, 6.21) |
|  | Male | Ref | Ref |
| **Marital Status** | Never married | 1.04 (0.82, 1.30) | 1.08 (0.82, 1.42) |
|  | Divorced/separated/widowed | 1.29 (0.74, 2.26) | 0.96 (0.51, 1.77) |
|  | Currently married | Ref | Ref |
| **Employment** | Unemployed | 1.65 (1.30, 2.10)** | 1.16 (0.82, 1.66) |
|  | Currently employed | Ref | Ref |
| **Income<sup>†</sup>** | Per 1000 rupee increase | 1.00 (0.99, 1.01) | 1.00 (0.99, 1.01) |
| **HIV status at buprenorphine initiation** | HIV + | 0.87 (0.57, 1.32) | 1.16 (0.73, 1.85) |
|  | HIV - | Ref | Ref |
| **Region** | Northeast ICCs | 1.90 (1.47, 2.46)** | 2.56 (1.76, 3.72)** |
|  | North/Central/Northwest ICCs | Ref | Ref |
| **Frequency of Counseling** | Any | 0.25 (0.20, 0.32)** | 0.21 (0.16, 0.27)** |
|  | None | Ref | Ref |
| **Average maintenance dose** | <8 mg | 1.18 (0.89, 1.58) | 1.28 (0.92, 1.79) |
|  | ≥8 mg | Ref | Ref |

OR = Odds ratio; aOR = Adjusted odds ratio
<sup>‡</sup>Adjusted multivariable model including all variables in univariable analysis
<sup>†</sup>Scaled by 1000 rupees
\*p-value < 0.05
\*\*p-value < 0.01

**Figure 3.**
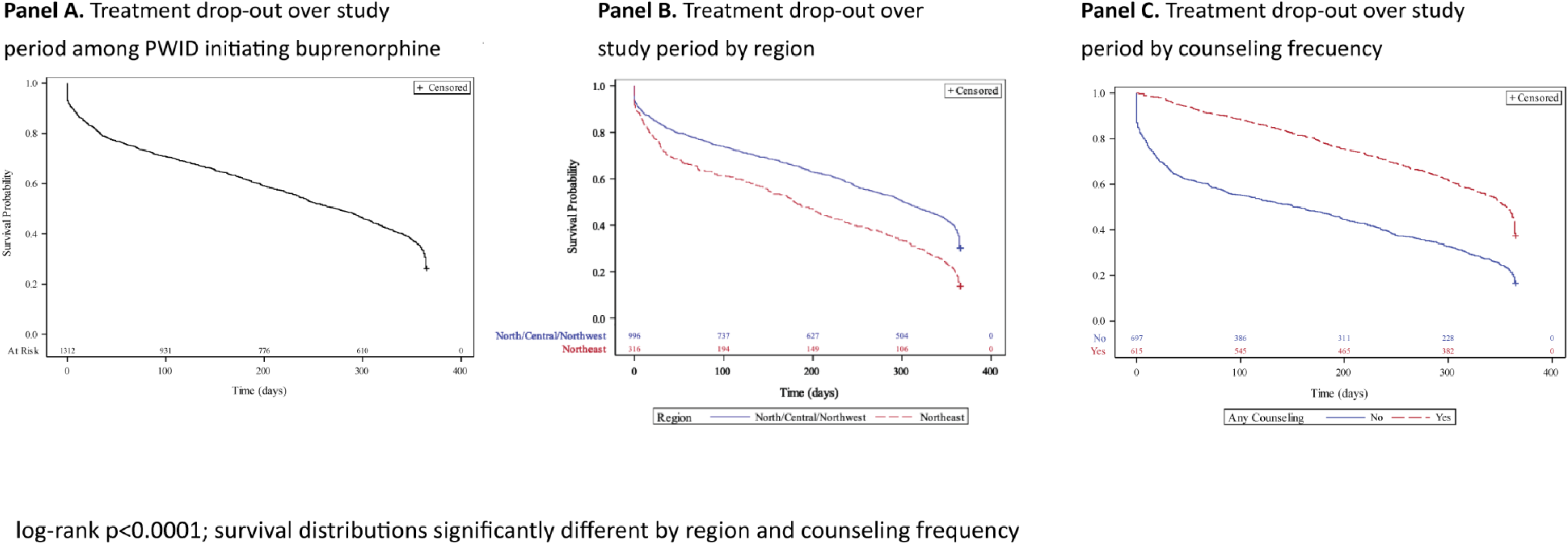
Kaplan Meir survival probabilities among PWID initiating buprenorphine at Integrated Care Centers (Panel A), by region (Panel B) and, by counseling frequency (Panel C)

### 1.3.6. Sensitivity analysis

As expected, the proportion of PWID experiencing a 30-day treatment interruption exceeded the proportion of PWID experiencing a 60-day treatment interruption (40% vs. 29%) and the likelihood of experiencing at least one treatment interruption remained higher among PWID at North/Central/Northwest ICCs compared to Northeast ICCs (NCC: 42%, NEC: 34%, p=0.0116). When comparing whether correlates in our univariable and multivariable models varied for treatment drop-out by 3 months versus 6 months, we found HIV-positive status associated with higher odds of drop-out by 3 months, but no other differences in results (Supplementary Table 1).

## 1.4 Discussion

Few studies have examined buprenorphine receipt and retention among PWID in public sector and/or public-private service delivery models in India. These studies been limited to five or fewer sites and/or a small number of PWID largely as part of operational research conducted in the initial roll-out of OST in the country^23,32–36^. In this study, by way of examining buprenorphine initiation, receipt and retention in more granular detail among PWID receiving services at ICCs across 7 cities in India, we constructed an opioid treatment cascade which points to significant programmatic and patient-level gaps as well as notable regional differences.

First, the opioid treatment cascade reveals a substantial gap in the provision of *any* substance use treatment to a sizeable proportion of PWID who receive services at ICCs. The absence of accurate size estimates of PWID in the cities where ICCs have been established^37^ mars an estimation of what proportion of the population at risk received OST at ICCs in 2018. However, among PWID who did visit ICCs and received at least one service in 2018, overall, 40% did not receive OST. One reason for this is that not all PWID may have met treatment criteria. However, given that the overwhelming majority who seek HIV testing services at ICCs report current injection drug use^38^, this gap may also represent missed opportunities to engage this population in substance use treatment. Another caveat to this observation is that ICCs do not explicitly record whether PWID may be receiving OST at other locations (e.g., at other NGOs or in the private sector). While this is a possibility in the Northeast cities (where there is greater availability of substance use treatment services), this is less likely a contributing factor in the North/Central/Northwest cities (where ICCs are among the very few available community-based public sector services for OST)^1,16,39^. Further, in our prior studies exploring patient perspectives regarding services at ICCs, PWID have indicated the availability of single-venue services as a significant draw, making it less likely that those receiving other services at ICCs would feel more inclined to receive OST at a different location^17,40^. Our data also suggests that there are no long delays between ICC registration and starting treatment. Instead, the fact that more than a third of PWID at ICCs are not receiving OST is likely explained by other factors that need to be studied further. One key structural factor that is universal across ICCs (and may explain the comparable proportion of PWID not on OST in both regions) is that OST dispensed to patients is procured from the government and there is a limited supply of medications and treatment slots (i.e., the demand far exceeds the supply). The need to expand OST considerably in the public sector (in terms of medication supply, treatment slots, and trained workforce) has been a long-standing policy recommendation^7,41,42^, which this study lends further support for.

Second, the retention outcomes at 6 months and 1 year suggests considerable attrition, particularly at the Northeast ICCs. Our findings are comparable to but also differ from findings reported in other studies conducted in India and elsewhere^23,31,33–36,43^. For example, overall retention, and regional retention at 6 months (even in the Northeast ICCs where retention is poorer) are comparable to that reported in other international studies^31,43^ and meet internal programmatic benchmarks of at least 50% retention in treatment at 6 months. However, while retention at 6 months among PWID at the North/Central/ Northwest ICCs in our study is comparable to the high retention reported in earlier studies in India, retention at the Northeast ICCs is considerably lower than that reported at two sites in the Northeast in a large study conducted more than a decade ago^23^. Additionally, overall retention of 35% at 1 year is also lower than that reported in older studies in India. This low retention at 1 year suggests that a significant proportion of PWID in both regions drop-out after 6 months.

Third, scrutinizing the quality of treatment engagement by evaluating average daily buprenorphine receipt and treatment interruptions paints a picture of poor engagement. Overall, only approximately 50% of PWID came more than two times to receive buprenorphine in the first 6 months following initiation. The median number of visits over 6 months was only 47 days. In other words, at least half of PWID have fewer than 1.5 months ‘worth of visits over a period of 6 months). Additionally, nearly a third experienced at least one treatment interruption over the course of treatment. In this context, the average maintenance dose that PWID received over the first 6 months following initiation is important to examine. The Indian OST guidelines^25^ for provision of buprenorphine recommend a lower daily maintenance dose of 8-12 mg compared to analogous guidelines in other countries^44,45^. While studies in many settings show that higher buprenorphine dose is associated with improved adherence and retention^46–51^, this recommendation in part was borne out of the earlier studies in India showing high retention with lower doses of buprenorphine^24,33–35,52,53^. However, even by the Indian OST guidelines, more than three quarter of PWID at ICCs received less than 8 mg of buprenorphine daily. Although the treatment of opioid use disorders is individualized, and doses are tailored to patient needs, this observation raises question about whether PWID at ICCs received adequate treatment (particularly since the maintenance dose did not change over the course of treatment for the majority). There is a need to understand reasons for providing much lower doses than that stipulated in guidelines (and whether these pertain to medication supply issues at ICCs versus other factors such a provider knowledge). In addition, few studies if any in India have evaluated the benefits of providing higher maintenance doses (above that recommended in the Indian OST guidelines) on adherence and retention.

In multivariable regression models, only region and receipt of counseling were significantly associated with treatment drop-out by 6 months. However, our prior qualitative study^40^ offers greater insights into the various structural and psycho-social barriers that PWID encounter for receipt of OST at ICCs. These barriers likely contribute to the sub-optimal engagement and retention observed in this study and have also been described in other studies in India^24,34,54^. For example, the policy of daily attendance to receive buprenorphine has been consistently highlighted as a significant barrier for adherence and retention (owing to a multitude of challenges such as lack of transportation, conflict with work and school schedules etc.)^34,40^. During the COVID-19 pandemic, recognizing care-seeking challenges during periods of uncontrolled transmission and lockdowns^55,56^, ICCs briefly shifted away from daily observed buprenorphine treatment to alternative prescribing schedules (such as providing a week or fortnight’s supply of medications at a time)^57^. Although ICCs have now reverted to daily observed treatment, there is a need to evaluate the impact of more convenient prescription schedules coupled with adequate dosing or longer acting buprenorphine formulations on retention and quality of engagement. Notably, in our study, over half of PWID did not have any counseling visits. That receipt of any counseling was associated with lower odds of treatment drop-out lends support that beyond merely making counseling services available at ICCs, a medication-assisted treatment model with structured evidence-based behavioral interventions needs to be actualized in practice. The regional differences we observed in our study are striking albeit somewhat surprising. Despite more evolved substance use and HIV services in cities in the Northeast, PWID at these ICCs not only had greater odds of treatment drop-out by 6 months, they also demonstrated poorer engagement across various measures of buprenorphine receipt compared to PWID at North/Central/Northwest cities. Additional research is needed to understand the contextual factors that influence these differences.

Our study has several limitations. Given its retrospective nature, we were limited to utilizing data that is collected and recorded as part of routine service delivery. This precluded examining other important factors that are not recorded (e.g., mental health co-morbidities, polysubstance use, family involvement and social support, incarceration etc.) but have been shown to influence engagement and retention in OST. ICCs also do not record if PWID who initiate OST are treatment naïve or have had prior history of treatment at other programs although for this study, we did ascertain that PWID who initiated buprenorphine in 2018 did not include those who had commenced treatment at ICCs and dropped-out in the preceding year but chose to re-initiate subsequently. This study was also underpowered to fully explore the impact of predictors such as higher buprenorphine dose and gender on retention. Specific to gender, less than 5% of PWID who receive services ICCs are cisgender women. While the vast majority of PWID in India are men, several studies, including our prior research in India have delineated the unique vulnerabilities and challenges that women who inject drugs experience for receipt of substance use treatment^36,40,58–60^. Although service delivery models exclusively for women have been piloted^61^, dedicated studies are needed to understand and ameliorate the gaps that women experience in buprenorphine treatment receipt and retention. Finally, grouping ICCs by region may have obscured site-specific differences, and findings are not generalizable to other cities.

Our study also has many strengths. As noted, studies in India examining buprenorphine receipt and retention are largely historical. This study therefore provides more contemporary estimates and has greater regional representation compared to prior studies. Additionally, biometric fingerprinting unlike other methods of documentation prone to gaps in recording allowed us to accurately capture daily visits, buprenorphine dose, as well as exact utilization of other services (e.g., counseling visits). Our findings of significant gaps in buprenorphine receipt and retention are particularly important in the context of data suggesting ongoing opioid use, continued risk behaviors and high HIV incidence among PWID receiving services at ICCs^38^. As the ICC model is under consideration for expansion across more cities in India, our findings can inform key changes needed in program delivery.

## 1.5 Conclusions

Taken together, the findings in our study point to the need to implement programmatic and patient-level interventions to improve initiation, engagement, and retention among PWID receiving buprenorphine at ICCs. Specifically, as opioid use disorders are chronic and relapsing, service delivery models such as ICCs that provide OST should aim to facilitate long term retention and improve quality of engagement in treatment by implementing cross-cutting patient-centered services adapted to regional contexts. Evidence suggests such an approach could provide benefits across several outcomes. While such interventions will require a paradigm shift in the approach to treatment of opioid use disorders in India’s public sector, they are urgently needed given the larger backdrop of burgeoning opioid injection epidemics across many cities.

## Data Availability

All data produced in the present study are available upon reasonable request to the authors.

## Acknowledgements

We thank the National AIDS Control Organization (NACO), India, all our partner non-governmental organizations throughout India, and patients at Integrated Care Centers, without whom this research would not have been possible.

## Funding

This study was supported by the National Institute on Drug Abuse of the National Institutes of Health (R01DA032059, R01DA041034, DP2DA040244 and K24DA035684). The study was also supported by the Harvard University Center for AIDS Research (CFAR), an NIH funded program (P30AI060354), The Johns Hopkins University CFAR (P30AI094189), The Elton John AIDS Foundation, and the Thrasher Research Fund. Its contents are solely the responsibility of the authors and do not necessarily represent the official views of the National Institutes of Health.

**Supplementary Table 1.** Predictors of treatment drop-out by 3 months among people who inject drugs initiating buprenorphine at Integrated Care Centers

|  |  | OR (95% CI) | aOR (95% CI)<sup>‡</sup> |
| --- | --- | --- | --- |
| **Age at buprenorphine initiation** | Per year | 1.00 (0.98, 1.02) | 1.00 (0.98, 1.02) |
| **Gender** | Female | 2.37 (1.03, 5.41)* | 2.14 (0.79, 5.80) |
|  | Male | Ref | Ref |
| **Marital Status** | Never married | 0.85 (0.66, 1.09) | 0.80 (0.59, 1.09) |
|  | Divorced/separated/widowed | 1.14 (0.63, 2.06) | 0.73 (0.37, 1.44) |
|  | Currently married | Ref | Ref |
| **Employment** | Unemployed | 1.59 (1.23, 2.05)** | 1.15 (0.78, 1.70) |
|  | Currently employed | Ref | Ref |
| **Income<sup>†</sup>** | Per 1000 rupee increase | 1.00 (0.99, 1.01) | 1.00 (0.99, 1.01) |
| **HIV status at buprenorphine initiation** | HIV + | 1.17 (0.76, 1.80) | 1.87 (1.12, 3.12)* |
|  | HIV - | Ref | Ref |
| **Region** | Northeast ICCs | 1.78 (1.36, 2.33)** | 2.96 (1.95, 4.49)** |
|  | North/Central/Northwest ICCs | Ref | Ref |
| **Frequency of Counseling** | Any | 0.15 (0.11, 0.20)** | 0.11 (0.08, 0.16)** |
|  | None | Ref | Ref |
| **Average maintenance dose** | <8 mg | 1.19 (0.86, 1.63) | 1.34 (0.92, 1.95) |
|  | ≥8 mg | Ref | Ref |

OR = Odds ratio; aOR = Adjusted odds ratio
\*Adjusted multivariable model including all variables in univariable analysis
<sup>†</sup>Scaled by 1000 rupees
\*p-value < 0.05
\*\*p-value < 0.01

